# Correlation of Sensory perception at different body parts with BMI using Two Point Discrimination

**DOI:** 10.1101/2023.03.13.23287193

**Authors:** Dev Himanshubhai Desai, Prahasth Dave, Neeraj Mahajan, Anita Verma

## Abstract

**Purpose:** Two-point discrimination (2PD) is the ability to discern that two nearby objects touching the skin are truly two distinct points, not one. It is often tested with two sharp points during a neurological examination and is assumed to reflect how finely innervated an area of skin is. objective was to find out the normal value of Two Point Discrimination on the Back, Finger, Forehead & Sole and to Correlate values of Two Point Discrimination on Back, Finger, Forehead & Sole with corresponding BMI according to Gender.

**Methods:** Two point discrimination test is a part of practical course of Physiology Department of the teaching institute. Here, the students perform the practical under strict supervision of professors, the data of the students’ practical is recorded for department’s personal use.

This data was collected with the permission from the head of the department and then analyzed to see the normal values of two-point discrimination and also to assess the correlation between Gender, BMI, & Two-point Discrimination(TPD). Pearson Correlation, T-test, SEM, CI95, Scatter diagrams were used to analyze and understand the data.

**Results:** Values of Two-Point-Discrimination(TPD) Average(in mm), SD, and Pearson Correlation(with BMI) at Back (Males=32.19 ± 4.94,r=0.808 & Females=29.38 ±5.29,r=0.848), Fingertips (Males=3.43 ± 1.52,r=0.0344 & Females=3.47 ± 1.52,r=0.0141), Forehead (Males=13.53 ± 2.36,r=0.0223 & Females=12.25 ± 2.61,r=0.198), Sole (Males=14.69 ± 3.54,r=0.00493 & Females=13.87 ± 3.36, r=0.0234)

**Conclusion:** The results depict the normal values of Two-point discrimination at the respective location. The normal values at a particular location in Males and Females are almost the same Strong Correlation of TPD value at Back with BMI can be justified as fat Accumulation increases in Skin with increasing BMI.

Fat mainly accumulates on the abdomen, thighs, and back and neck but not on Forehead, Fingertips & soles.

Fat accumulation hypothetically leads to increase skin surface leading to virtually variable per area relative decreased skin innervation resulting in higher TPD values and Strong Positive Correlation.

TPD values at Forehead, Fingertips & soles do not correlate with BMI.

## Introduction

Nervous system has evolved from very primitive beings to humans. It used to be a diffuse net of neurons in lower animals (1) which evolved to become a complex multi-neuronal central (2) and peripherally(3) distributed system in humans. The need of it to save the organism and show the way to food has increased to complex calculation and sensory-motor output system.

One of the two parts of the peripherally located part of the nervous system is Sensory input (4)acceptance and conduction to the central nervous system for interpretation and necessary reaction. The inputs are taken by many different nerve endings and are transmitted to the spinal cord. One of the main input signal is called touch and that can be divided in two types of sensation namely Fine touch (Tract of Gall and Bardach in spinal cord) (5) and Crude touch (anterolateral tract). (5)

Touch sensation allows one to have ability to tell two different points. This is essentially important when the whole area of skin is innervated by a single nerve-root (the area is called a Dermatome) (6). Touch sensations allows us to differentiate two different points even in a same dermatome but that has a limit. Innervation by a single nerve-root leads to overlapping(7) and hence, a threshold distance is formed where if the points are wider apart then the threshold, subject can appreciate the two points otherwise even though there are two points externally stimulated, subject only feels one. Distance in every human is different but there is a general trend found that it is highest on back and very low on fingertips. (8)

Test to measure this threshold distance is called Two-point discrimination test. Two-point discrimination (2PD) is the ability to discern that two nearby objects touching the skin are truly two distinct points, not one. It is often tested with two sharp points during a neurological examination and is assumed to reflect how finely innervated an area of skin is. (8) and instrument used for this is called a two pointed compass aesthesiometer. (9) (10) This device has two hands with pointed (non-harmful) tip and an Arc distance measure tape attached at the joint.

Among diagnostic tests, 2PD assessment seems to be particularly important because it is often used in clinical practice to evaluate the severity of peripheral nerve injuries and to monitor recovery and response to treatment. (11)

The uses of it are important, as in assessing the prospects for surgical and other kinds of rehabilitation in tetraplegia and stroke patients the two-point discrimination test, correctly performed by experienced examiners, is of great value (12) as in a study, Thirty adults with completed ischemic stroke and left paresis participated in the study. The ability for 2PD sensation was significantly reduced in all examined fingers on the affected side in comparison to the non-affected side(13)

Two-point discrimination test can be used from checking nerve damage in a patient to check the effect of fat deposition on relative nerve innervation density.

## Methodology

A cross sectional study was designed to collect data. Students studying at Smt. NHL Municipal Medical College were enrolled as study participants. Physiology Department of the teaches this as their regular practical curriculum and records the data for departments personal use. Data is recorded under the strict observation of Professors and authors. The data of the Static tow point discrimination test was collected from the department.

### Static two point discrimination test:- (8)

The two point aesthesiometer is take and set at 0(so that both tips are touching each other) and put on the predetermined location on the subject’s body; while putting this points, the subject is asked to call out how may points it can appreciate. The distance between the tips is gradually increased and two tips are simultaneously put on the subject’s body every time and asked how many point are appreciable. This process is repeated till the subject calls out that it can now appreciate two points. At this time, the measurement on the arc measure tape id recorded and the test on that body part is completed.

All the data gathered was digitalized and was used to analyze the outcomes.

### Study design: - Cross sectional record based Study

Study population: - after excluding students by the exclusion criteria, total of 222 students were enrolled from which 64 were males and 158 were female students. All of them were of the age group of 17-19.

### Inclusion criteria

▪ Student who were willing to give consent for the test.
▪ Students with no skin condition or nerve defects.

### Exclusion criteria

∘ Students who were not willing to consent.
∘ Students already diagnosed with any skin conditions.
∘ Students already diagnosed with peripheral neuropathy
∘ Student with type 1 Diabetes
∘ Students who were sick or felt under the weather.

Static two-point discriminatory test was conducted at following body parts of students to attain the data

· Back
· Forehead
· Sole
· Palm

Statistical analysis done with

· Microsoft excel
· SPSS 20

Statistical tests conducted

· Average
· Standard deviation
· Comparative analysis across ranges
· Standard Error of Mean (SEM)
· Confidence interval (CI95)
· Pearson correlation (R value)
· Statistical significant test (unpaired t test) (p value)
· Analysis of variance (ANOVA) (f statistics)
· Charts and tables

Data will be made available to journal and editors upon request from the author and we would be very happy to share it.

## Results

Demography (General details about population): -

1. Gender (Table 1)

2. Age
  ∘ All data from Participants were gathered when they entered the Medical college as First year students and hence all were from 17-19 age group.
3. Two-point discrimination threshold values at different body parts in Males. (Table 2)

|  | FEMALES | MALES | TOTAL |
| --- | --- | --- | --- |
| Number of participants | 158 | 64 | 222 |

4. Two-point discrimination threshold values at different body parts in Females. (Table 3)

3. Two-point discrimination threshold values at different body parts in Males. (Table 2)
| Males | Back | Forehead | Fingertip | Sole | Anova (F-Statistics)(p-values) |
| --- | --- | --- | --- | --- | --- |
| Average | 32.188 | 13.531 | 3.430 | 14.688 | 0.0001 |
| Standard Deviation | 4.944 | 2.364 | 1.519 | 3.541 |  |
| SEM | 0.618 | 0.295 | 0.190 | 0.443 |  |
| CI5 | 1.211 | 0.579 | 0.372 | 0.867 |  |

5. Unpaired t test values (p-value) at different body parts between Males and Females (Table 4)

4. Two-point discrimination threshold values at different body parts in Females. (Table 3)
|  | Back | Forehead | Fingertip | Sole | Anova (F-Statistics)(p-values) |
| --- | --- | --- | --- | --- | --- |
| Average | 29.380 | 12.253 | 3.475 | 13.873 | 0.0001 |
| Standard Deviation | 5.292 | 2.611 | 1.516 | 3.359 |  |
| SEM | 0.421 | 0.208 | 0.121 | 0.267 |  |
| CI5 | 0.825 | 0.407 | 0.236 | 0.524 |  |

6. Pearson Correlation coefficient value (R-value) showing correlation between two-point discrimination threshold value at given body part against BMI, Height and Weight in males. (Table 5)

**5. Unpaired t test values (p-value) at different body parts between Males and Females (Table 4)**
| Males vs Females | Back | Forehead | Fingertip | Sole |
| --- | --- | --- | --- | --- |
| P value | 0.0003 | 0.0008 | 0.838 | 0.1074 |

7. Pearson Correlation coefficient value (R-value) showing correlation between two-point discrimination threshold value at given body part against BMI, Height and Weight in Females. (Table 6)

| Male (for All BMI ranges) | Back | Forehead | Fingertip | Sole |
| --- | --- | --- | --- | --- |
| BMI | 0.808 | 0.022 | 0.034 | 0.005 |
| height | -0.163 | -0.032 | 0.116 | 0.182 |
| Weight | 0.718 | -0.016 | 0.095 | 0.137 |

| Female (for All BMI ranges) | Back | Forehead | Fingertip | Sole |
| --- | --- | --- | --- | --- |
| BMI | 0.848 | 0.198 | 0.014 | 0.023 |
| height | -0.028 | 0.019 | 0.035 | -0.157 |
| Weight | 0.764 | 0.197 | 0.022 | -0.047 |

## Discussion

1. With the help of Table 2, these points can be denoted for Two points discrimination in males.
  a. Back has the highest amount of threshold distance followed by Sole and forehead having a narrow margin in between. The lowest amount of discrimination value can be seen at fingertip.
2. Looking at Table 3, it is evident here that
  a. Same as in males, high amount of threshold difference can be found at back followed sole and forehead having a narrow margin. The lowest threshold can be found at fingertip.
3. Figure 1,2,3,4 are sufficient here to show that females have a better sensory acuity and low threshold distance to discriminate between two points.
  a. At back, along with forehead and sole, females show significantly lower threshold distance than in males when it comes to discriminating two points apart.
  b. At fingertip, the average of two-point discrimination distance is almost similar in between males and females.
  c. This can be supported by Table 4 where we can see the results of unpaired t-test in between male and female participants showing that the difference between males and females at back and forehead is statistically significant, whereas at sole and fingertip the difference is not statistically significant
4. Table 5 with Figure 5,6,7,8 is evident is showing that in males
  a. At back the correlation between discrimination distance is highly associated with the BMI and weight and negative and weakly associated with height.
  b. A similar weak correlation is evident with all weight, height and BMI is visible when compared with two-point discrimination value at Fingertip, sole or forehead.
5. Table 6 with Figure 9,10,11,12 shows the similar trend in females in comparison of BMI
  a. At back, BMI and weight is positively strongly correlated with two-point discrimination, but height is negatively and weekly associated
  b. And when it comes to fingertip, sole and forehead, the correlation is weak.

- The result depicted here are pretty consistent with the results found in previous research papers at back, forehead, sole and fingertip.
  ∘ In a book called Bates’ guide to Physical Examination and History Taking (14) similar amount of discrimination threshold distance is found. The same results are mentioned by Wikipedia (8)
  ∘ A other research paper about 2PD values at fingertip also found the same results (15)
- Literature supports our finding of seeing a smaller discrimination distance value and more two-point discriminatory power in females then in males.
  ∘ In a research paper by Louis et al., Statistical analysis of the data revealed the following: female subjects consistently tended to discriminate at shorter distances when compared with male subjects at corresponding sites. (16)

➢ Our hypothesis behind the result: - c
  ∘ With increasing BMI, the major factor of the increase is weight gain. Higher weight means a higher deposition of fat cells around abdomen and back as central adiposity is major fat storage system in weight gain. (17)
  ∘ With increased weight, the adiposity stretches the skin and the total surface are of the body part increases. (18)
  ∘ This increase in total surface area is not aided with increase in peripheral nerve innervation.
  ∘ Hence, new innervation of skin doesn’t occur and the nerve fibers are stretched only to innervate the increase of skin area.
  ∘ This leads to. Which in turns is represented as increase in the two-point discrimination.

**Figure 1.**
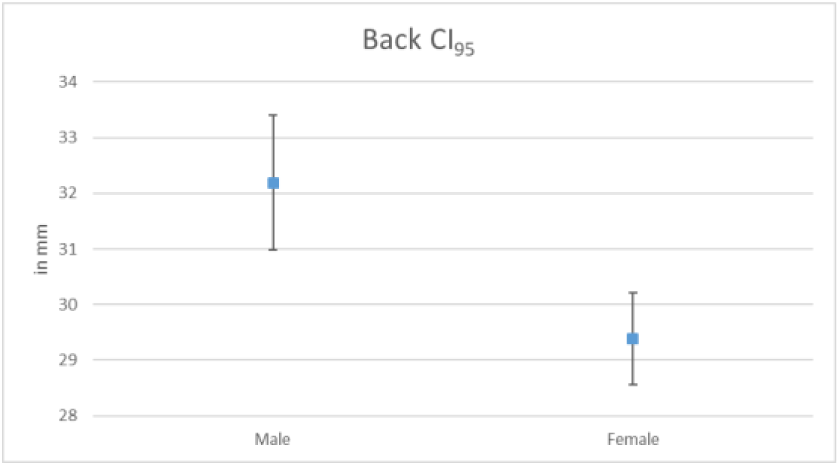

**Figure 2.**
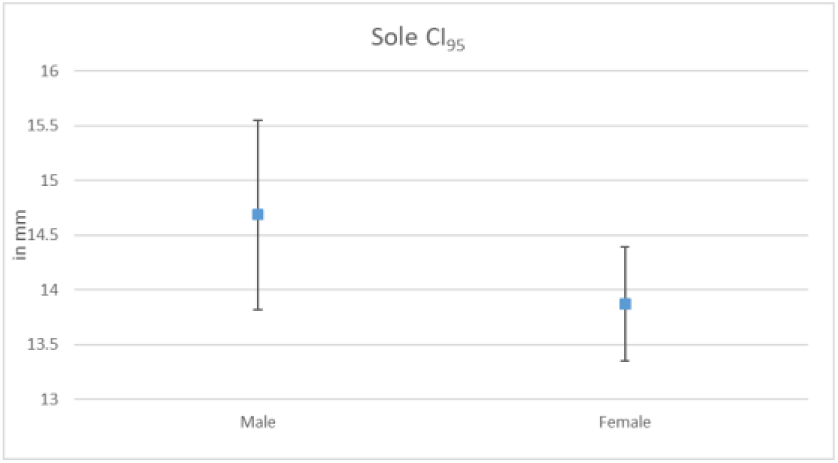

**Figure 3.**
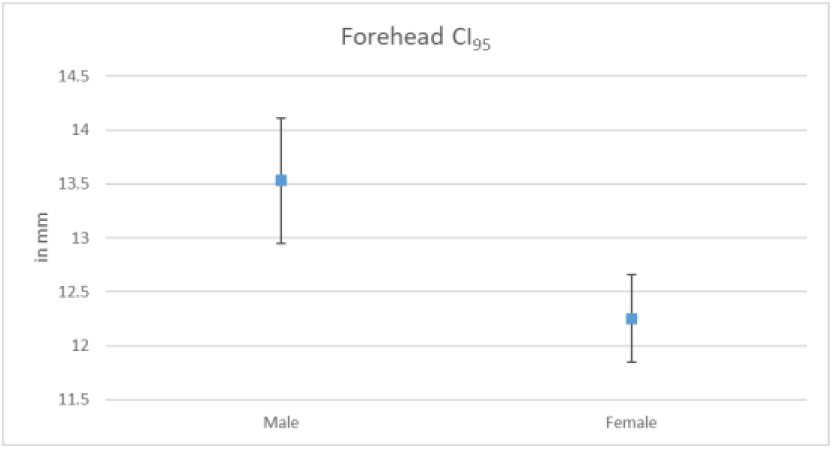

**Figure 4.**
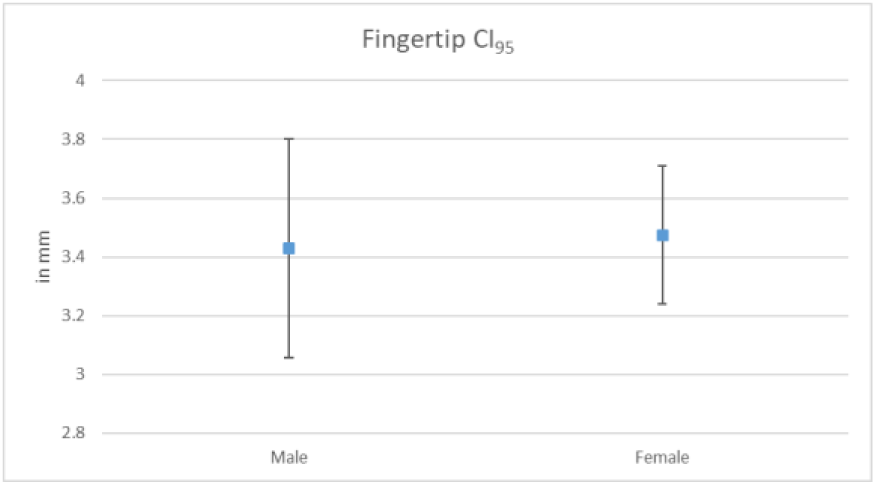

**Figure 5.**
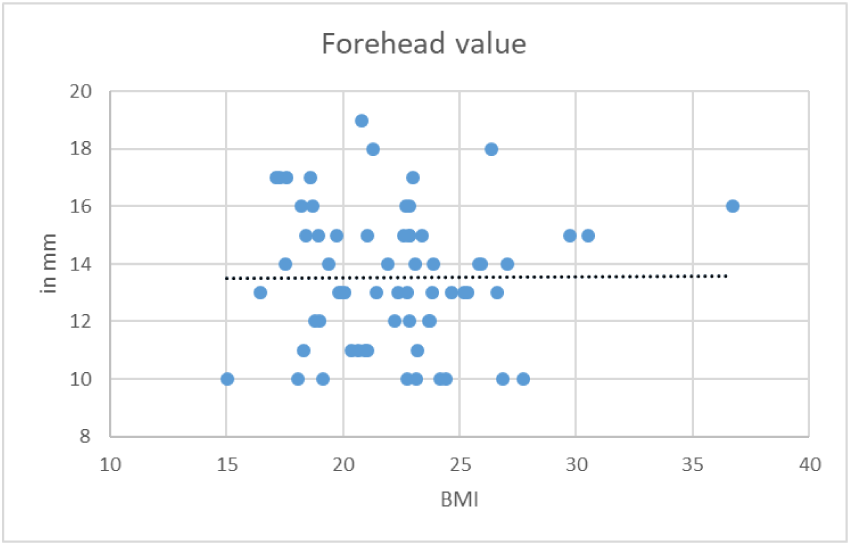

**Figure 6.**
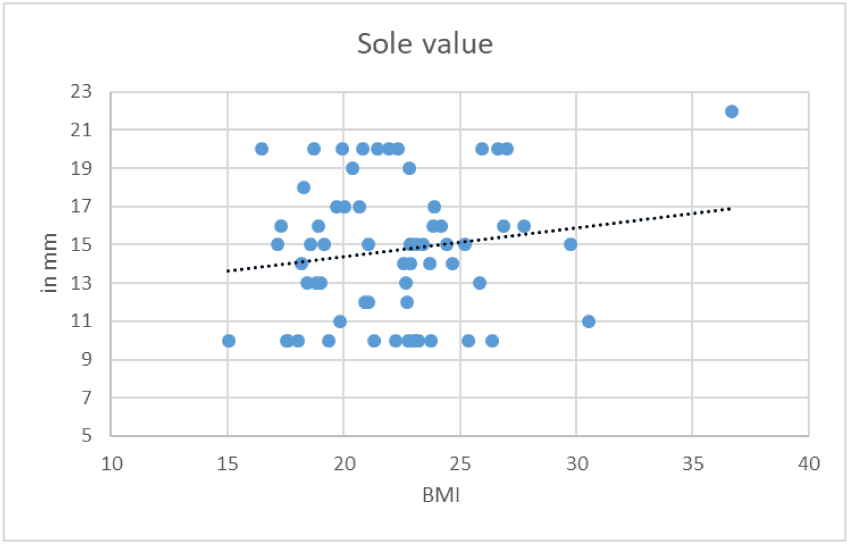

**Figure 7.**
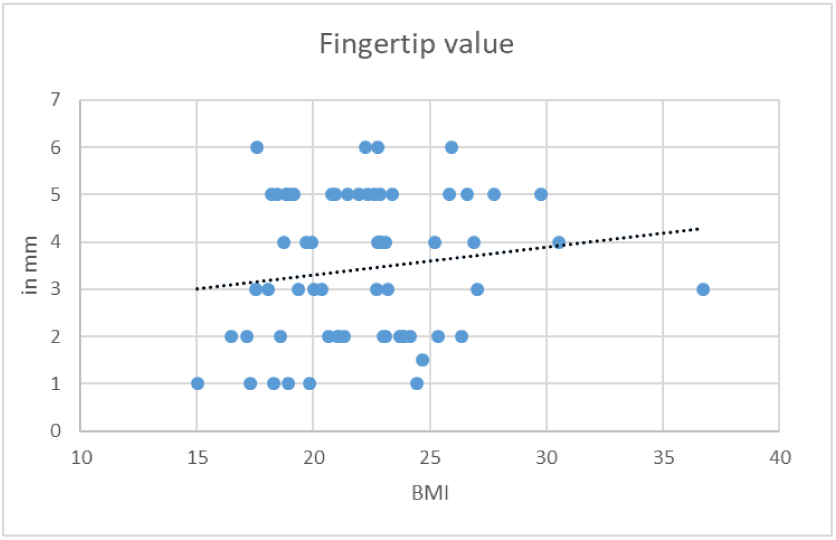

**Figure 8.**
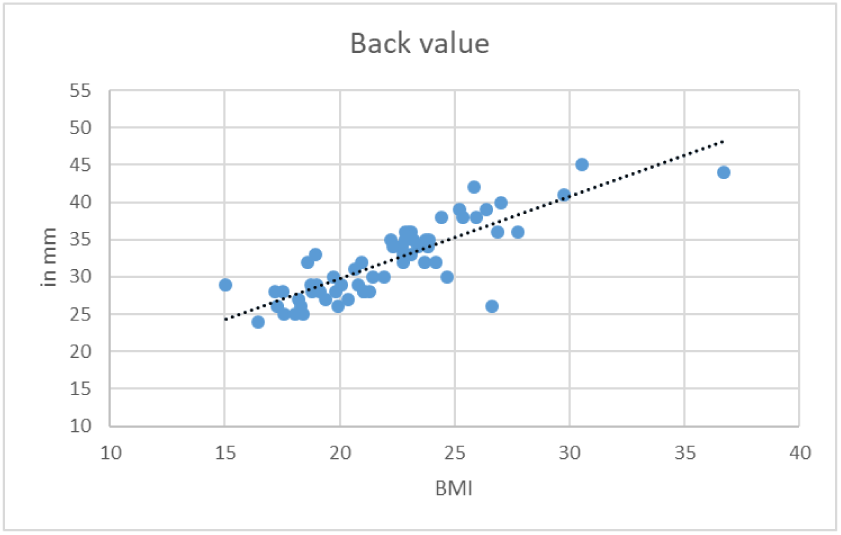

**Figure 9.**
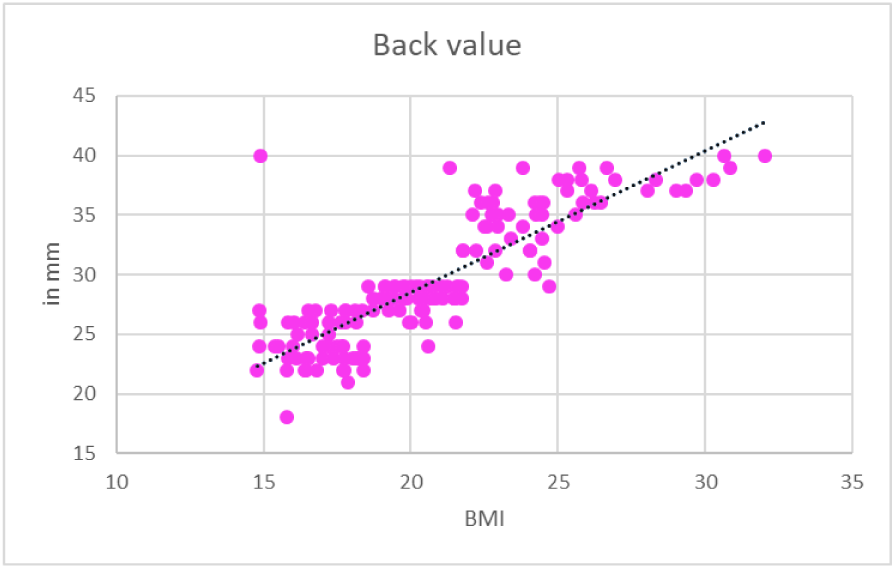

**Figure 10.**
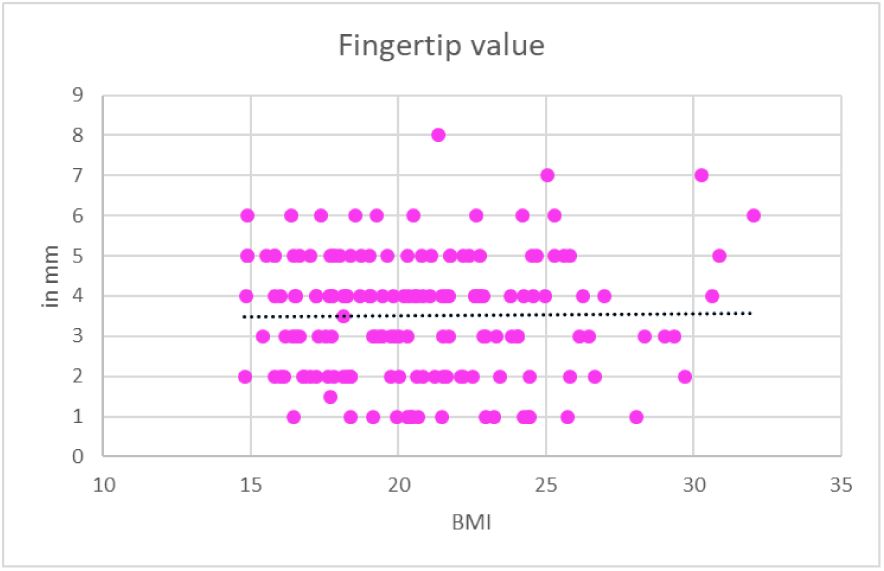

**Figure 11.**
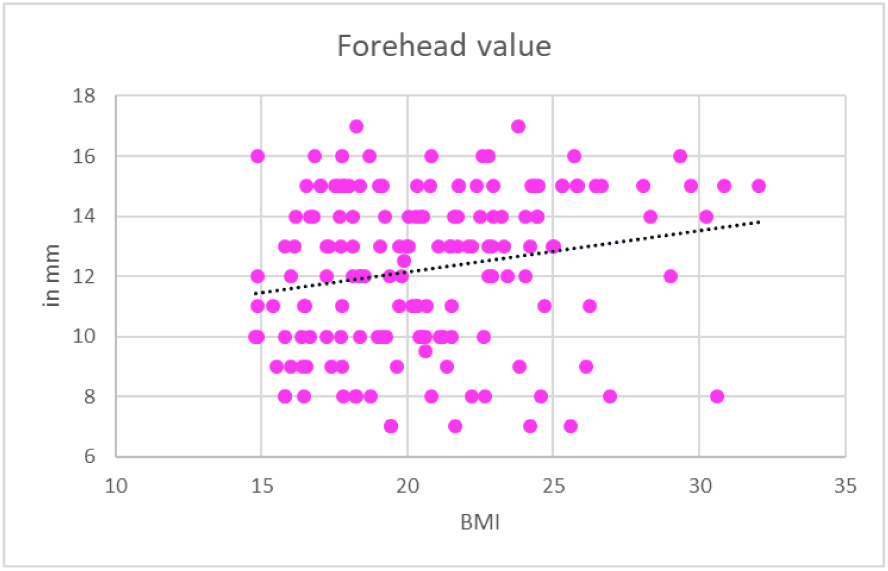

**Figure 12.**
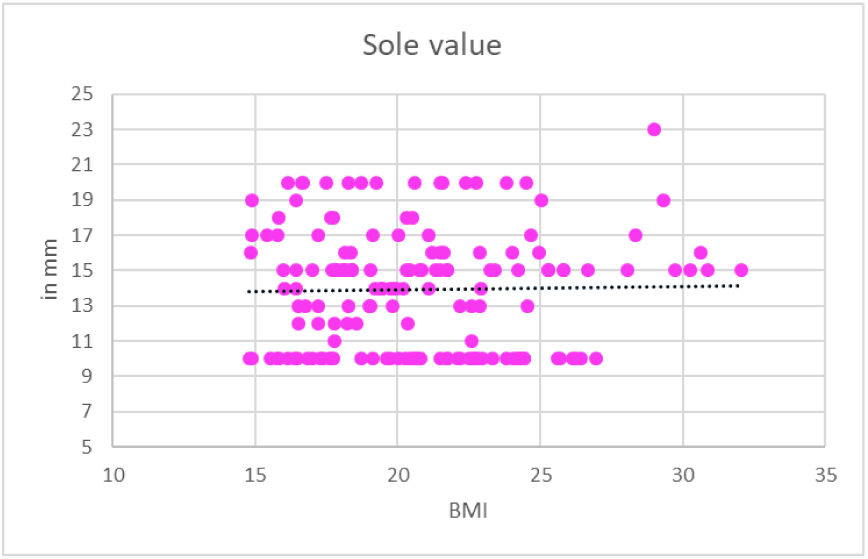

## Conclusion

It is evident with the tables that threshold of to discriminate two points in the same dermatome is highest at back followed by forehead and sole. The least threshold distance can be found at fingertips as it is one of the most important tactile organ in the body.

it was found in the results that female have significant higher discriminatory power (meaning they have a lower threshold distance and can discriminate two points when they are lesser apart then in males) then males in all the aspects except on fingertips where no significant difference in the discriminatory power was found.

This also shows that in both males and females Two-point discrimination values at back are strongly correlatable with BMI but it is weakly correlatable with two-point discrimination values at forehead, sole and fingertip (even weak negative correlation coefficient values were also found)

## Limitations and for future

➢ Same study with higher sample size would yield a more statistically stable results. \
➢ The hypothesis that has been shown needs a targeted review and research with high funding and resources to understand the effect of obesity better on skin and more importantly how the skin is innervated.

## Data Availability

All data produced in the present study are available upon reasonable request to the authors

## Declarations

### Funding

The authors have received no funding or have any financial gain from this study

### Conflict of interest

The authors have no conflict of interest to disclose

## Statements and Declarations

The authors have no relevant financial or non-financial interests to disclose. The authors did not receive support from any organization for the submitted work.

The Authors have no conflict of interest

## Notes

### Competing Interest Statement

The authors have declared no competing interest.

### Funding Statement

This study did not receive any funding

### Author Declarations

IRB os Smt. NHLMMC (Shrimati Nathiba Hargovindas Lakhamichand Municipal Medical College) gave Ethical approval for this work

